# Glioma genetic profiles associated with electrophysiologic hyperexcitability

**DOI:** 10.1101/2023.02.22.23285841

**Authors:** Steven Tobochnik, Maria Kristina C. Dorotan, Hia S. Ghosh, Emily Lapinskas, Jayne Vogelzang, David A. Reardon, Keith L. Ligon, Wenya Linda Bi, Stelios M. Smirnakis, Jong Woo Lee

## Abstract

Distinct genetic alterations determine glioma aggressiveness, however the diversity of somatic mutations contributing to peritumoral hyperexcitability and seizures is uncertain. In a large cohort of patients with sequenced gliomas (n=1716), we used discriminant analysis models to identify somatic mutation variants associated with electrographic hyperexcitability in a subset with continuous EEG recording (n=206). Overall tumor mutational burdens were similar between patients with and without hyperexcitability. A cross-validated model trained exclusively on somatic mutations classified the presence or absence of hyperexcitability with an overall accuracy of 70.9%, and improved estimates of hyperexcitability and anti-seizure medication failure in multivariate analysis incorporating traditional demographic factors and tumor molecular classifications. Somatic mutation variants of interest were also over-represented in patients with hyperexcitability compared to internal and external reference cohorts. These findings implicate diverse mutations in cancer genes associated with the development of hyperexcitability and response to treatment.

## INTRODUCTION

Seizures are a common comorbidity in patients with brain tumors, leading to disability and decreased quality of life.^1,2^ However, identifying which patients are at risk and implementing targeted treatment strategies remains a clinical challenge. Seizures have been consistently recognized at high rates in low-grade glial and glioneuronal tumors, even as tumor classification has evolved dramatically over the last several decades.^3,4^ Advances in tumor genomic analysis have led to significant changes in the subtyping of glial tumors according to their molecular profiles, while the integration of histologic and molecular diagnoses has improved clinical predictions of glioma aggressiveness and survival outcomes.^5−7^ Certain molecular pathways can also facilitate the interaction of glioma cells with adjacent neurons and regulate the development of peritumoral electrophysiologic hyperexcitability, which may further promote tumor progression and impact patient survival.^8,9^ Yet, the relationship between seizures at glioma diagnosis and electrographic hyperexcitability remains poorly understood. The high rates of seizure-freedom after tumor resection suggest that distinct mechanisms may promote seizures at glioma diagnosis or recurrence compared to cases with uncontrolled seizures and enduring electrographic hyperexcitability.^10^

The contribution of glioma genetic variants to electrographic hyperexcitability and seizures has been characterized in only a subset of isolated somatic mutations to date.^11−16^ These effects have been demonstrated at the level of specific variants such that different activating somatic mutations in the same gene can have unique mechanisms and variable influences on epileptogenesis.^12^ We hypothesize that diverse glioma somatic mutations facilitate peritumoral hyperexcitability and that genetic profiling may allow for better identification of patients at risk of clinically relevant hyperexcitability than current models incorporating traditional risk factors. Here we analyze the distribution of glioma somatic mutations in patients with and without electrographic hyperexcitability measured by continuous electroencephalography (cEEG) and use statistical classification models to identify putative variants associated with hyperexcitability (Figure 1).

**Figure 1.**
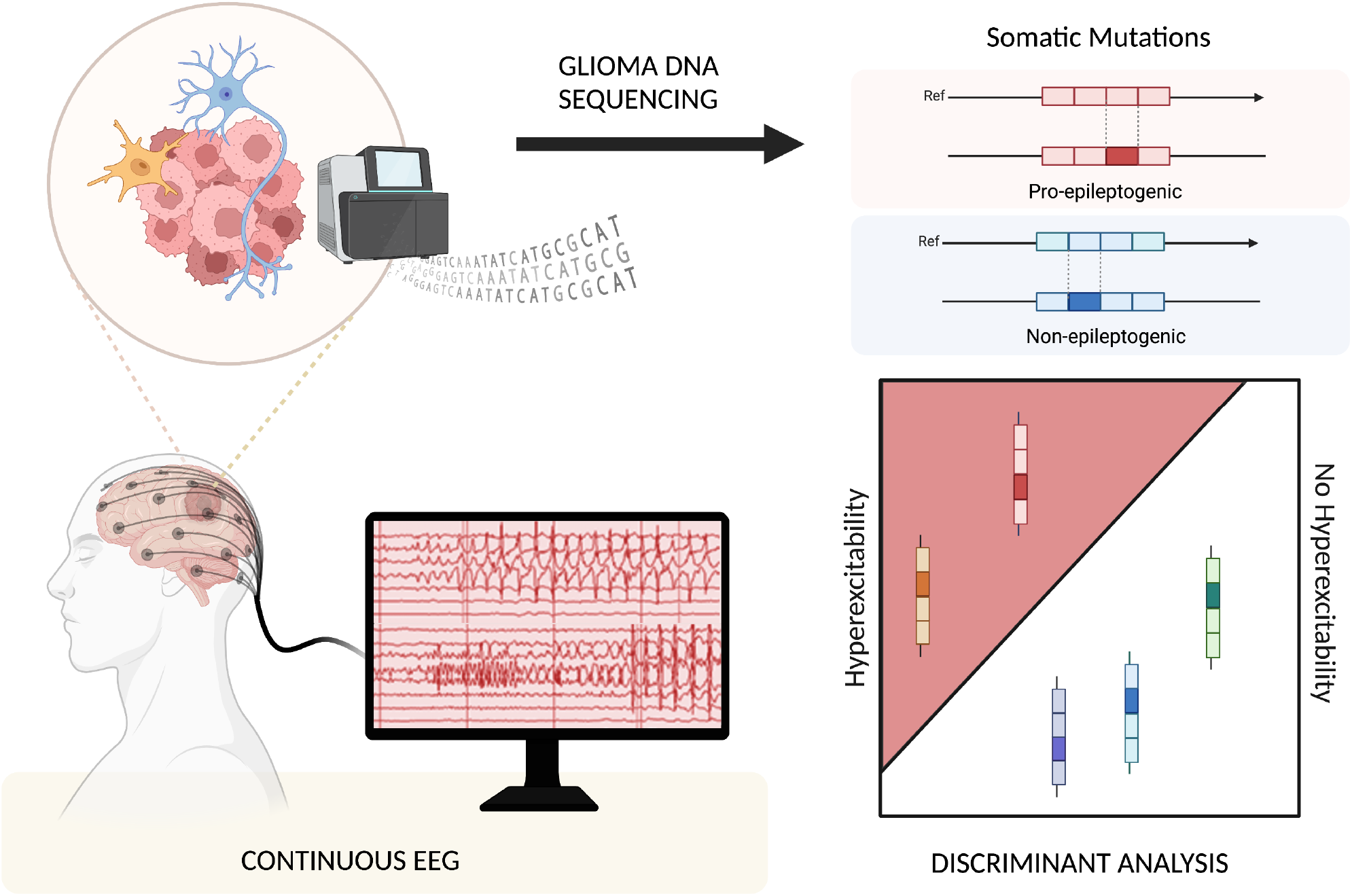
Study design scheme. Overview of the study design in a large cohort of patients with sequenced gliomas for somatic mutation variants, some of which were hypothesized to be pro-epileptogenic. Continuous electroencephalography (EEG) in a sub-cohort was used to define electrographic hyperexcitability. Discriminant analysis was applied for classification of somatic mutation variants associated with hyperexcitability. Created with BioRender.com.

## METHODS

### Study Design

The total cohort included all adult patients (18 years and older) with an integrated pathologic diagnosis of WHO grade 1-4 glioma who underwent tumor genetic testing with OncoPanel between 2013-2022 at Dana-Farber Cancer Institute (DFCI). Oncopanel, a targeted gene next generation sequencing assay developed and validated at the Center for Cancer Genome Discovery (DFCI) and the Center for Advanced Molecular Diagnosis (Brigham and Women’s Hospital), detects single nucleotide variants, insertions, and deletions as well as copy number variants, and intronic regions of rearrangement.^17,18^ Cases were excluded if there was insufficient tissue sampling for OncoPanel testing. Associated clinical oncologic data were extracted from an integrated prospective oncology database (OncDRS).^19^ A phenotyping cohort for the primary analysis included the subset of patients who underwent continuous video EEG monitoring (Figure 1), identified through cross-referencing with the validated Critical Care EEG (CCEEG) database at Brigham and Women’s Hospital.^20^

All protocols for Human Subjects Research were approved with waiver of informed consent by the Institutional Review Boards of the Dana-Farber/Harvard Cancer Center (DF/HCC #21-425) and VA Boston Healthcare System (#1710792-1).

### Clinical Electrophysiology and Seizure Outcomes

Continuous EEG (cEEG) was performed using XLTEK systems (Natus Medical Inc, Pleasanton, CA) as clinically indicated. The duration of monitoring was determined at the discretion of the treating medical team. All cEEG recordings were included for each patient and designated as either pre-operative or post-operative relative to their first tumor surgery. Hyperexcitability was defined as the presence of lateralized periodic discharges and/or focal electrographic seizures (LPD/SZ) according to American Clinical Neurophysiology Society (ACNS) Standardized Critical Care EEG Terminology.^21^ As a secondary outcome, two levels of seizure control were designated based on patients’ anti-seizure medication requirement at the latest clinical encounter, stratified as ≤1 medication or ≥2 medications to achieve seizure control.

### Tumor Genetic Sequencing

Tumor specimens were analyzed for somatic mutation variants using targeted exome OncoPanel testing for approximately 500 cancer-associated genes as previously described.^22^ Briefly, genomic DNA was extracted, fragmented for library preparation, amplified by PCR, followed by hybridization capture and Illumina HiSeq 2500 sequencing. The standard minimum tumor purity for analysis was 20%. Variants in non-coding regions and variants with <50x coverage or a variant allele fraction (VAF) <10% were excluded during data preprocessing based on prior validation studies.^17^

### Genetic Profile Classification

After cross-referencing with electrophysiology data, variants with at least two occurrences were retained for the final analysis. To evaluate for clustering and collinearity between somatic mutation variants, a pairwise correlation matrix was constructed. Significant correlations were identified using the Benjamini-Hochberg procedure to control the false discovery rate (FDR) at a level of α=0.05.

A regularized discriminant analysis (RDA) model was applied for dimensionality reduction using the klaR package.^23,24^ Regularization was used to mitigate the effects of multicollinearity arising from correlated variants,^23,25^ which limited linear discriminant analysis. RDA classified the binary outcome of hyperexcitability based exclusively on somatic mutation variant inputs. Tuning and variable selection with leave-one-out cross-validation was performed in a bidirectional stepwise manner using a stop criterion of improvement in correctness rate <0.25%. To test whether the established relationship between the canonical *IDH1-R132H* variant and clinical seizures would also translate to electrographic hyperexcitability, two RDA models were evaluated: one with *IDH1-R132H* designated the starting variable (M_IDH_R132H_), and the other at baseline with no specified starting variable (M_BL_). RDA posterior probabilities were used to calculate the area under the receiver operating characteristic curves (AUC) and bootstrap resampling performed to estimate 95% confidence intervals (2000 replicates).^26^

### Functional Analysis

The functional consequences of missense variants discovered in M_BL_ and M_IDH_R132H_ (excluding *IDH1-R132H*) were predicted using PolyPhen-2.^27^ Gene set enrichment analysis was performed using g:Profiler to evaluate potential molecular mechanisms, biologic pathways, and shared disease processes associated with variants identified by the classification models.^28^ Mechanistic enrichment analysis included the molecular function, cellular component, and biological process domains of Gene Ontology (Gene Ontology Consortium).^29,30^ Phenotypic enrichment analysis was performed using the Human Phenotype Ontology (Monarch Initiative). For each analysis, inferred electronic annotations without manual or curator review were excluded and a term size limit of 1000 was applied. The Benjamini-Hochberg procedure was used to control the FDR at a level of α=0.05.

### Statistical Analysis

Comparisons between categorical and continuous variables were performed using two-sided Fisher Exact and Mann-Whitney U tests, respectively. Multivariate logistic regression was used to evaluate the association of genetic profiles to hyperexcitability and seizure control outcomes relative to other clinical predictors, including age, sex, WHO tumor classification, and tumor localization. The incidence of somatic mutation variants in gliomas with hyperexcitability were compared to two reference cohorts representing the general glioma population with a baseline level of hyperexcitability. An internal reference cohort consisted of patients who did not undergo cEEG recording. Patients with glioma and somatic mutation data in The Cancer Genome Atlas (TCGA-GBM and TCGA-LGG) composed an external reference cohort. Binomial tests were used to compare incidence rates with the Benjamini-Hochberg procedure to control the FDR at a level of α=0.05. Data processing, model development, and statistical analysis was performed using R 4.0.3 (The R Foundation for Statistical Computing).

## RESULTS

### Clinical Cohort Characteristics

We analyzed 1716 adult patients with sequenced gliomas (WHO grade 1-4), of whom 206 (12.0%) had cEEG performed. Demographics and tumor pathologies were similar between patients with and without cEEG (Table 1). Within the cEEG cohort, hyperexcitability defined by the presence of lateralized periodic discharges and/or focal electrographic seizures (LPD/SZ), was present in 82/206 (39.8%) patients. Patients with hyperexcitability had greater cEEG recording days (mean 2.9 vs 1.5 days, p=1.46e-14, Table 2) and were more likely to be treated with multiple anti-seizure medications (OR=2.67, 95%CI 1.40-5.15, p=0.002). EEG focality aligned with tumor localization, with temporal lobe involvement most frequently observed in both patients with and without hyperexcitability, followed by frontal and parietal lobe involvement (Table 2).

**Table 1.** Demographic and oncologic characteristics of patients with and without cEEG.

|  | Total<br>N=1716 | With cEEG<br>N=206 | Without cEEG<br>N=1510 |
| --- | --- | --- | --- |
| Age, mean years (range) | 53.2 (18-94) | 55.6 (22-86) | 52.9 (18-94) |
| Sex, female N (%) | 748 (43.6) | 75 (36.4) | 673 (44.6) |
| WHO classification, N (%) |  |  |  |
| Glioblastoma | 779 (45.4) | 95 (46.1) | 684 (45.3) |
| Diffuse astrocytoma, IDH-WT, NEC | 335 (19.5) | 49 (23.8) | 286 (18.9) |
| Astrocytoma, IDH mutant | 343 (20.0) | 36 (17.5) | 307 (20.3) |
| Oligodendroglioma, IDH mutant | 173 (10.1) | 17 (8.3) | 156 (10.3) |
| Pediatric-type diffuse HGG | 30 (1.7) | 4 (1.9) | 26 (1.7) |
| Pediatric-type diffuse LGG | 56 (3.3) | 5 (2.4) | 51 (3.4) |
| WHO grade, N (%) |  |  |  |
| 1 | 21 (1.2) | 1 (0.5) | 20 (1.3) |
| 2 | 201 (11.7) | 22 (10.7) | 179 (11.9) |
| 3 | 181 (10.5) | 20 (9.7) | 161 (10.7) |
| 4 | 1252 (73.0) | 158 (76.7) | 1094 (72.5) |
| Unspecified | 61 (3.6) | 5 (2.4) | 56 (3.7) |
| Disease status at biopsy, N (%) |  |  |  |
| Primary | 1362 (79.4) | 164 (79.6) | 1198 (79.3) |
| Recurrent | 236 (13.8) | 31 (15.0) | 205 (13.6) |
| Unspecified | 118 (6.9) | 11 (5.3) | 107 (7.1) |
| OS, median days | 674 | 516 | 706 |

cEEG, continuous electroencephalography; HGG, high grade glioma; IDH, isocitrate dehydrogenase;
LGG, low grade glioma; NEC, not elsewhere classified; OS, overall survival; WHO, World Health
Organization, WT, wildtype

**Table 2.**
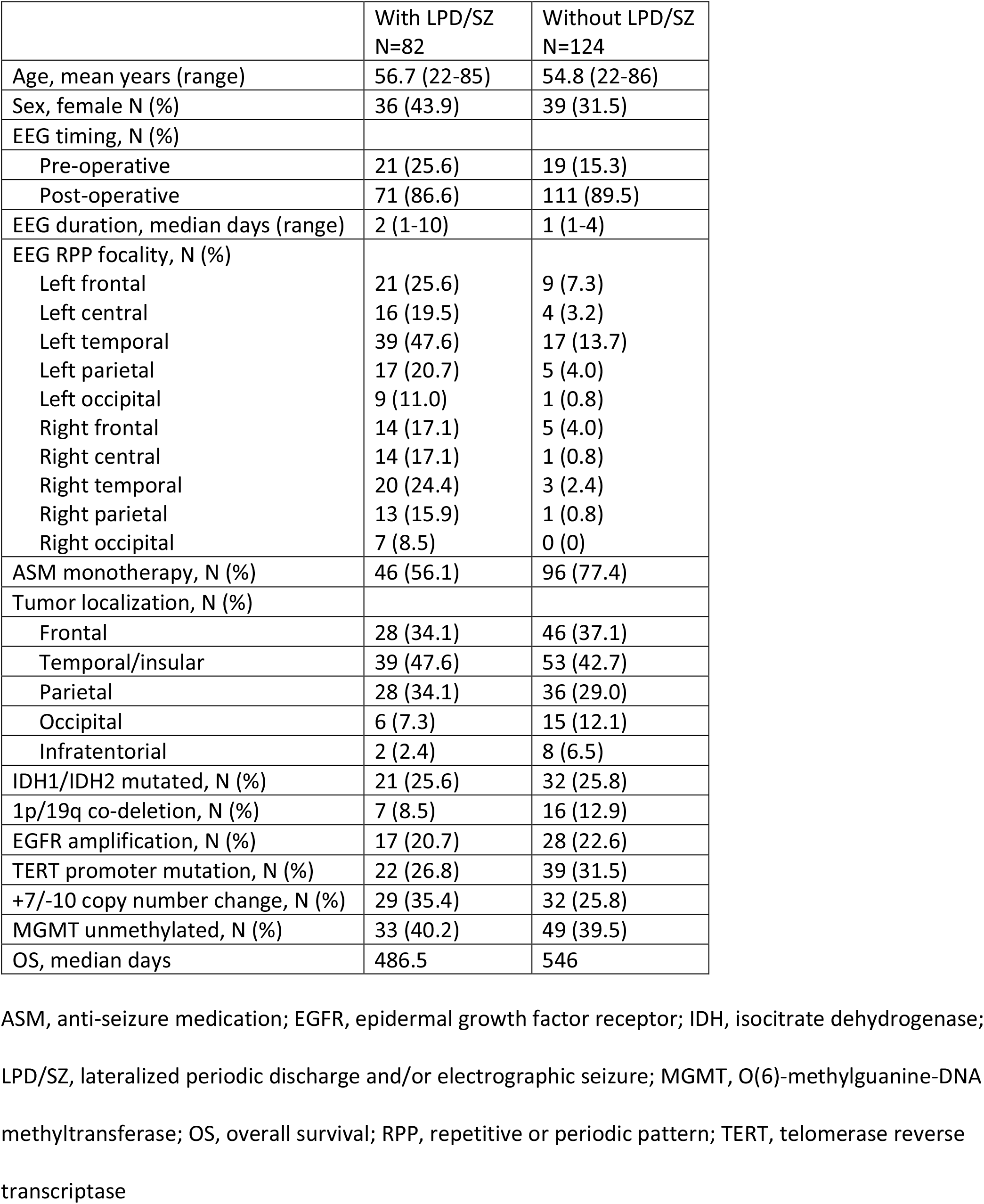
Clinical characteristics of patients with and without glioma-related hyperexcitability.

### Distribution of Glioma Somatic Mutations

Tumor specimens were analyzed for somatic mutation variants in a panel of cancer-associated genes using targeted exome next-generation capture sequencing (DFCI OncoPanel). After preprocessing, 11261 unique somatic mutation variants were detected in 508 different genes across the total cohort, including 1055 variants with multiple occurrences. The most common variant was the canonical *IDH1-R132H* mutation, occurring in 444/1716 (25.9%) patients (Figure 2a). In the cEEG cohort, 1412 unique somatic mutation variants were detected across 387 genes, including 73 variants in 38 genes with multiple occurrences. Pairwise correlation with Benjamini-Hochberg correction was performed to assess for collinearity between variants in the cEEG cohort (Figure 2f). Highly correlated variants were identified among 184/2628 (7.0%) pairs involving 61 genes (Figure 2g).

**Figure 2.**
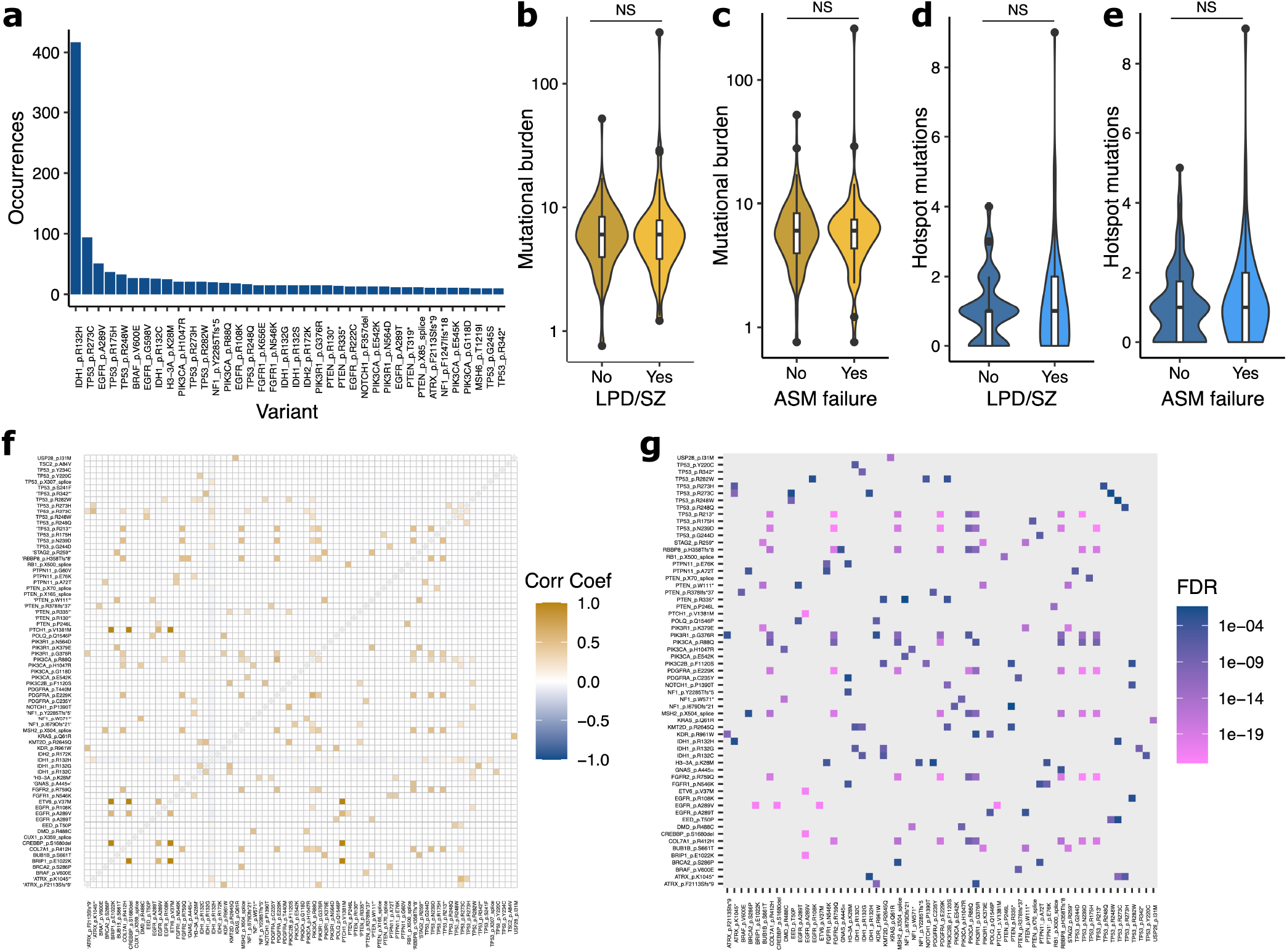
Distribution and correlation of glioma somatic mutations in the cohort. (a) Skew of somatic mutations with variants occurring at least 10 times shown here, representing 0.3% of all unique variants. (b-e) Comparison of tumor mutational burden (b,c) and hotspot variant clustering (d,e) in association with lateralized periodic discharges and/or electrographic seizures (LPD/SZ) or anti-seizure medication (ASM) failure. (f) Pairwise correlation matrix of variants with >1 occurrence in the EEG cohort. (g) Significantly correlated variant pairs adjusted by false discovery rate (FDR).

### Extent of Genetic Variation and Hyperexcitability

We used several metrics to evaluate whether the extent of glioma genetic heterogeneity was associated with hyperexcitability. Given the inclusion of multiple glioma subtypes and grades, we assessed characteristic genetic alterations associated with the WHO tumor classification for adult-type diffuse gliomas.^6^ The frequency of these genetic alterations were similar between patients with versus without hyperexcitability (*IDH1/IDH2* mutated 25.6 vs 25.8%, p=1.0; 1p/19q co-deletion 8.5 vs 12.9%, p=0.55; *EGFR* amplification 20.7 vs 22.6%, p=0.86; *TERT* promoter mutation 26.8 vs 31.5%, p=0.53; chromosome +7/-10 copy number alteration 35.4 vs 25.8%, p=0.16, respectively).

We used tumor mutational burden (TMB) indices to capture the accumulation of somatic mutations at the individual level. TMB was similar between patients with versus without hyperexcitability (mean 9.6 vs 6.6 mutations per Mb, p=0.64, Figure 2b). Additionally, the proportion of cases with hyperexcitability were comparable between hypermutated (≥10 mutations per Mb) and non-hypermutated (<10 mutations per Mb) tumors (0.48 vs 0.39, OR 1.42, 95%CI 0.51-3.91, p=0.49). There were no significant differences in TMB (mean 10.3 vs 6.7 mutations per Mb, p=0.96) between patients requiring ≥2 anti-seizure medications or ≤1 anti-seizure medication for seizure control (Figure 2c).

For hotspot variants defined by multiple occurrences in the cEEG cohort, we analyzed the rates of co-occurrence of different variants per individual. The frequency of hotspot variants was similar in patients with and without hyperexcitability (mean 1.1 vs 1.0, p=0.83, Figure 2d), as well as in patients requiring ≥2 anti-seizure medications or ≤1 anti-seizure medication for seizure control (mean 1.2 vs 1.0, p=0.42, Figure 2e).

### Identification of Somatic Mutation Variants Associated with Hyperexcitability

We evaluated two cross-validated regularized discriminant analysis (RDA) models to classify the presence or absence of hyperexcitability with stepwise variable selection, one with the canonical *IDH1-R132H* variant as the starting variable (M_IDH_R132H_), and the other at baseline with no specified starting variable (M_BL_). M_IDH_R132H_ achieved an overall accuracy of 70.9% with incorporation of 15 variants and AUC 0.66 (95%CI 0.59-0.72, Figure 3c). M_BL_ achieved an overall accuracy of 70.9% with incorporation of 14 variants and AUC 0.64 (95%CI 0.59-0.70, Figure 3d). There were 13 variants in common between the two models (Figure 3a-b), occurring in *ATRX, DMD, KRAS, TP53, EGFR, PIK3CA, and USP28*.

**Figure 3.**
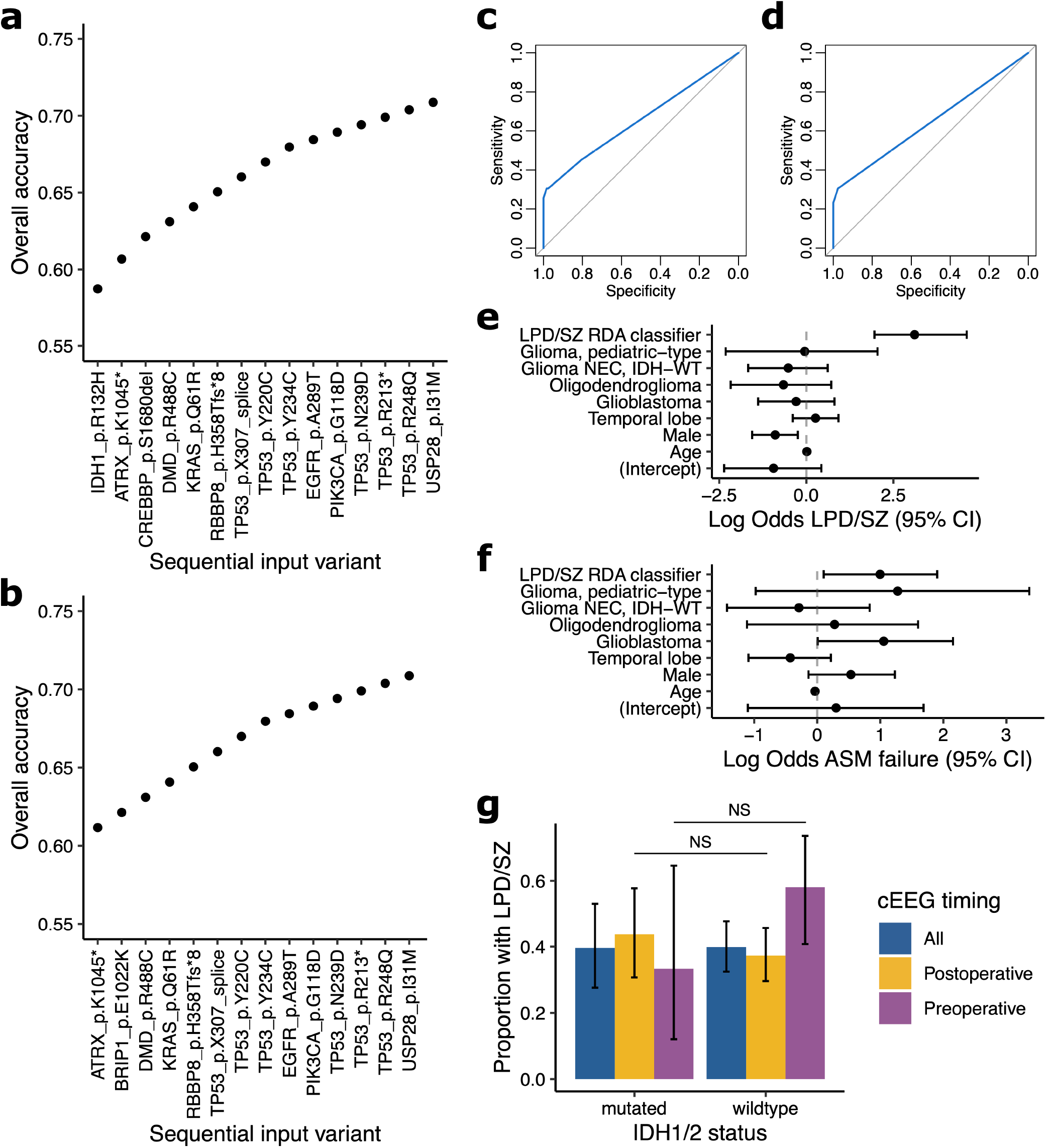
Somatic mutation profiles associated with hyperexcitability. (a-b) Stepwise variable selection using regularized discriminant analysis (RDA) and leave-one-out cross-validation for classification of presence or absence of hyperexcitability, with IDH1-R132H as the starting variable (a) or unsupervised (b). (c-d) Area under the receiver operating characteristic curves for models in (a) and (b), respectively. (e-f) Multivariate logistic regression forest plots of hyperexcitability (e) and ASM failure (f) estimates including hyperexcitability classifications by the RDA model in (b), WHO tumor classification, temporal lobe involvement, and demographic predictors. (g) Hyperexcitability rates compared between IDH-wildtype and IDH-mutated gliomas, stratified by timing of continuous EEG (cEEG) relative to the initial surgery.

In composite, the variant profile identified by M_BL_ was associated with the presence of hyperexcitability (OR 17.4, 95%CI 5.02-94.0, p=8.41e-9) as well as the secondary clinical outcome of anti-seizure medication failure (OR 2.55, 95%CI 1.04-6.23, p=0.027). To confirm this effect was not due to other variants correlated with the M_BL_ profile, we identified the set of positively correlated non-intersecting variants (n=16) and found no significant relationship with either hyperexcitability (OR 1.26, 95%CI 0.67-2.36, p=0.45) or anti-seizure medication failure (OR 1.17, 95%CI 0.60-2.27, p=0.635). To assess the association of the M_BL_ variant profile with hyperexcitability while controlling for standard clinical seizure risk factors, the hyperexcitability classifications were included in multivariate logistic regression analysis including age, sex, WHO classification, and temporal/insular lobe tumor localization. The M_BL_ classifier was a predictor for both the outcome of hyperexcitability (β=3.11, SE=0.66, p=2.14e-6, Figure 3e) as well as anti-seizure medication failure (β=1.00, SE=0.46, p=0.029, Figure 3f).

Given the established association of *IDH1* and *IDH2* mutations with pre-operative clinical seizures and prolonged survival,^5,15,31^ we further explored the contribution of *IDH* mutation status on hyperexcitability stratified by the timing of cEEG relative to initial tumor surgery (Figure 3g). The proportion of cases with hyperexcitability were similar between patients with or without any *IDH* variant regardless of the timing of cEEG (pre-operatively 0.33 vs 0.58, p=0.18; post-operatively 0.44 vs 0.37, p=0.37).

### Validation of Pro-Hyperexcitability Somatic Mutation Variants

To support the hypothesis that a specific somatic mutation profile may be over-represented in gliomas with hyperexcitability defined by LPD/SZ, we compared the incidence of hotspot variants in the hyperexcitability cohort with two reference cohorts representing the general glioma population with a baseline level of hyperexcitability with or without clinical seizures. In comparison with an internal reference cohort of patients without cEEG (n=1510), there were 16 variants in unique genes over-represented in the hyperexcitability cohort (Benjamini-Hochberg adjusted false discovery rate (FDR) <0.05, Figure 4a). In comparison to an external The Cancer Genome Atlas (TCGA) reference cohort (n=887), 25 variants in 23 unique genes were over-represented (FDR <0.05), including all the variants with increased incidence relative to the internal reference cohort (Figure 4b). Of the 15 unique variants identified by M_IDH_R132H_ and M_BL_ (excluding *IDH1-R132H*), 8/15 (53.3%) and 10/15 (66.7%) were observed at increased rates compared to the internal and external reference cohorts, respectively.

**Figure 4.**
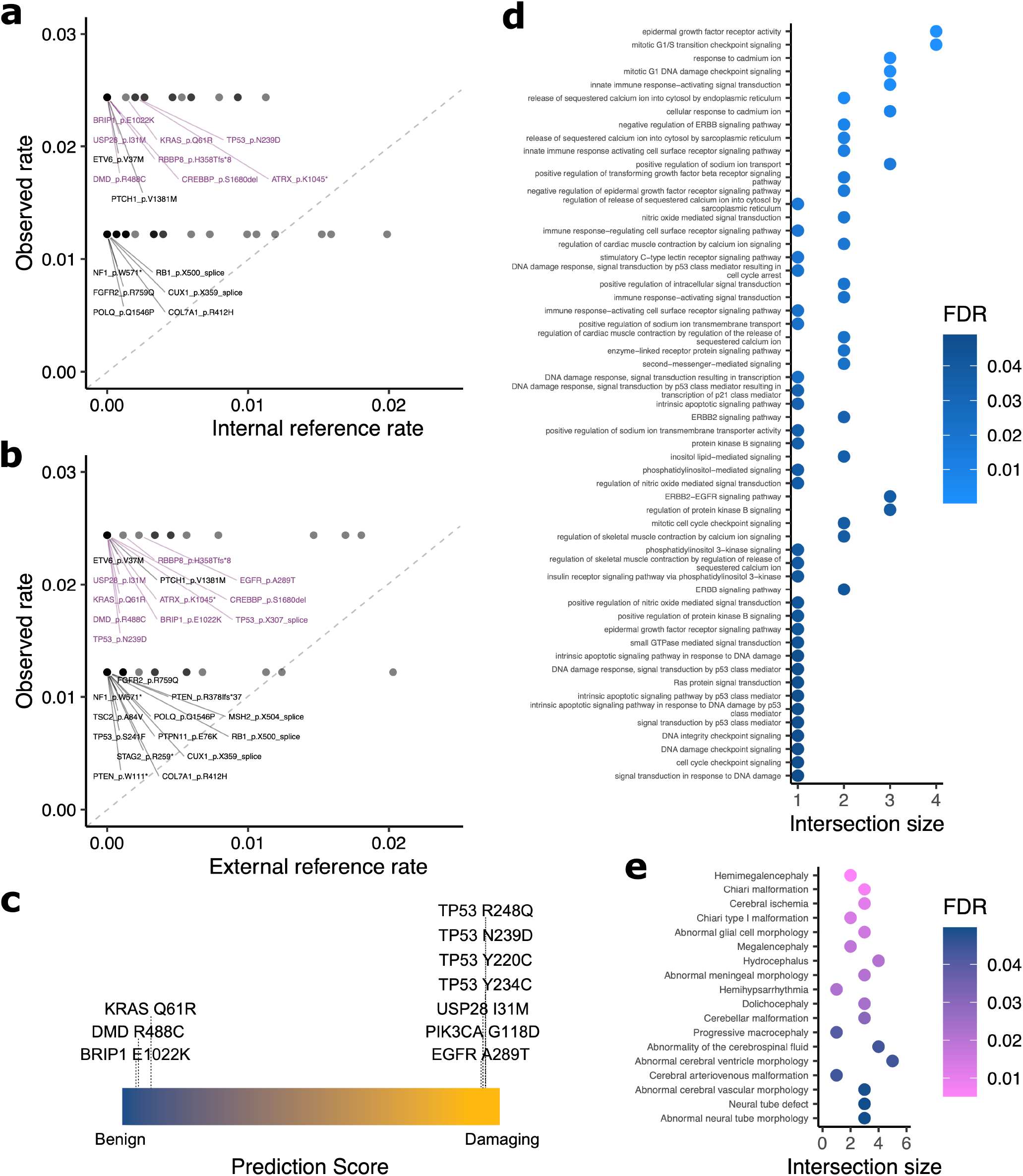
Hyperexcitability variant validation and gene set enrichment. (a-b) Quantile-quantile plot of somatic mutation rates in the hyperexcitability cohort in reference to the internal non-EEG cohort (a) and external TCGA cohort (b). All labeled variants were significantly over-represented (FDR <0.05) in the hyperexcitability cohort, and those in purple were also identified by the discriminant analysis models. (c) Polyphen-2 prediction scores for missense variants of interest. (d) Functional enrichment of the discriminant analysis gene set in cell signaling, receptor activation, and ion regulation processes in the Gene Ontology. (e) Functional enrichment of the discriminant analysis gene set in brain malformations, non-neoplastic lesions, and epilepsy syndromes in the Human Phenotype Ontology.

### Functional Enrichment Analysis

The functional consequences of missense variants in the M_BL_ and M_IDH_R132H_ mutation profiles (excluding *IDH1-R132H*) were predicted using PolyPhen-2, indicating a damaging effect for the *TP53, USP28, PIK3CA*, and *EGFR* variants, and a benign effect for *KRAS, DMD*, and *BRIP1* variants (Figure 4c). To evaluate molecular function and biological processes associated with the M_BL_ and M_IDH_R132H_ gene set, we performed enrichment analysis using the Gene Ontology. As expected, there was a predominant over-representation of cell cycle regulation, nucleic acid processing, and gene expression signaling pathways. Among the 377 enriched processes (FDR <0.05), we identified 46 (12.2%) specifically involved in cell signaling, 10 (2.7%) in receptor activation, and 12 (3.2%) in ion regulation (Figure 4d). Similarly, the majority of phenotypic enrichment using the Human Phenotype Ontology involved neoplastic diseases across organ systems. One epilepsy syndrome (hemihypsarrhythmia) was identified among the 521 enriched phenotypes (FDR <0.05). Brain malformations and non-neoplastic lesions made up 17/521 (3.3%) phenotypes (Figure 4e) and cranial malformations an additional 14/521 (2.7%) phenotypes.

## DISCUSSION

In this cohort of patients with glioma and tumor somatic sequencing, we used statistical classification methods to identify somatic mutation profiles associated with EEG hyperexcitability. We found that specific variants were enriched compared to internal and external reference cohorts and improved estimates of hyperexcitability in models incorporating traditional demographic factors and tumor molecular classifications. The results indicate that diverse somatic mutations in cancer-associated genes, but not the overall extent of tumor genetic variation, may promote epileptogenicity.

The definition of hyperexcitability used here encompassed standardized electroclinical measures of cortical irritability and has been previously validated as an independent predictor of mortality in *IDH*-wildtype diffuse gliomas.^21,32^ Lateralized periodic discharges and electrographic seizures represent an extreme of the hyperexcitability spectrum while aligning with common metrics used in animal models, typically seizures or electrophysiologic ictal-like events.^12,33,34^ The similar rates of LPD/SZ in pre-operative and post-operative cEEGs and the correlation with degree of long-term seizure control support the notion that this type of hyperexcitability is an enduring property of certain gliomas, which may involve different mechanisms than clinical seizures at tumor presentation. This is a likely reason for the lack of increased epileptogenicity seen with *IDH* mutations here, as *IDH*-mutated gliomas have predominately been associated with pre-operative seizures.^15,31^ However, since the majority of cEEG recordings were performed post-operatively, the study was not sufficiently powered to evaluate pre-operative hyperexcitability.

The OncoPanel gene set was developed based on known cancer associations rather than epileptogenic processes, yet many of these genes are implicated in a range of pathologies. Alterations in growth factor signaling and cell cycle regulation, predominately involving the PI3K-AKT-mTOR pathway,^35^ have been associated with both neoplasia and non-neoplastic cortical malformation syndromes.^36−39^ Somatic mutations in genes overlapping with some of the hyperexcitable variants identified in this study (*PIK3CA, KRAS, PTEN, NF1, TSC2*) have been reported in association with focal cortical dysplasia and drug-resistant epilepsy.^40^ Indeed, the hyperexcitable gene set here demonstrated phenotypic enrichment of multiple brain malformations, suggesting the potential for similar epileptogenic mechanisms mediated by somatic mutations in gliomas. However, whether these genetic alterations contribute to epileptogenicity by the same mechanisms in different cell types is uncertain.

Gliomas exhibit high inter- and intra-tumoral genetic variability.^41−44^ While we did not detect an effect of the extent of genetic variation on hyperexcitability, the potential for polygenic influences cannot be excluded. It is likely that different distributions of somatic mutations exist in other glioma cohorts, and thus the specific pro-epileptogenic profiles may vary as well. Furthermore, the vast majority of somatic mutations were single occurrences, presenting both a computational challenge and limiting genotype-phenotype association. We only included somatic mutations with multiple occurrences in the models here to avoid falsely attributing isolated variants as either pro-epileptogenic or non-epileptogenic.

This analysis was limited to the oncologic genetic data available. Further investigations with whole genome tumor sequencing, gene expression data, and molecular function analysis are needed to elucidate underlying mechanisms of these mutation profiles at an individual variant level, as well as the contribution of somatic mutations in non-cancer genes. The associations identified are also specific to the definition of hyperexcitability used here. Given that cEEG was performed for clinical suspicion of seizures and the majority of the cohort did not have hyperexcitability, under-detection in the cEEG cohort was likely low, however the baseline rates of hyperexcitability in the internal and external reference cohorts are uncertain.

In summary, glioma genetic profiling can identify diverse somatic mutations in cancer genes associated with peritumoral hyperexcitability and response to treatment. Further characterization of the epileptogenic mechanisms and interactions between variants will support targeted management strategies for glioma-related epilepsy.

## Data Availability

All data produced in the present study are available upon reasonable request to the authors.

## ACKNOWLEDGEMENTS

This research was supported by the Department of Veterans Affairs, Veterans Health Administration, V1CDA2022-68 (ST) and NIH/NINDS R03-NS091864 (JWL). The authors would like to acknowledge the Dana-Farber Cancer Institute Oncology Data Retrieval System (OncDRS) for the aggregation, management, and delivery of the clinical and operational research data used in this project. The content is solely the responsibility of the authors. Additional reference data used here was generated by the TCGA Research Network: https://www.cancer.gov/tcga. The authors thank Dr. Ellie Pavlick for her comments on the manuscript.

